# From survival to empowerment: A PLS-SEM analysis of residential aging-suitability for empty-nest seniors in urban China

**DOI:** 10.64898/2026.03.24.26349222

**Authors:** Xiaokang Liu, Yiming Peng, Yalong Xing, Hong Li

**Author notes:** Corresponding authors: (YLX) (HL).

## Abstract

The rapid aging of the population in urban China has led to a significant increase in empty-nest households, necessitating a rigorous evaluation of residential environment suitability. Grounded in Person-Environment Fit theory, this study develops and validates a multidimensional Aging-Suitability Index (ASI) specifically for urban empty-nest seniors. We analyzed survey data from 753 participants across 19 provinces using Partial Least Squares Structural Equation Modeling (PLS-SEM). The comprehensive structural model demonstrated robust explanatory power (*R²* = 0.754). The results reveal a hierarchical mechanism of needs: safety features and physical design serve as the survival foundation, exerting the most substantial direct effects on overall suitability. Accessibility was found to enhance suitability primarily by fostering perceived independence, indicating a psychological mechanism of empowerment (Variance Accounted For = 67.35%). Furthermore, intelligent technology and social support function as complementary resources that improve the environment-person fit. These findings suggest that aging-in-place interventions should prioritize mandatory safety upgrades while integrating accessibility modifications to sustain functional autonomy for independent seniors.

## Introduction

The rapid demographic transition in urban China has precipitated a fundamental structural change in household compositions, characterized by the exponential growth of “empty-nest” seniors—older adults residing without co-resident children [1–3]. As traditional multigenerational family support systems erode under the pressures of urbanization, empty-nest seniors face heightened vulnerability to physical decline, cognitive impairment, and psychological distress [4–7]. For this specific demographic, the concept of “aging in place” is no longer merely a preference for familiar surroundings [8]; rather, the residential environment itself assumes the critical role of a primary caregiving proxy [9, 10]. Consequently, accurately assessing and enhancing the “aging-suitability” of urban residential spaces has emerged as an urgent public health and urban planning imperative [11, 12].

Extensive epidemiological and gerontological evidence underscores that suboptimal physical environments significantly exacerbate health risks for older adults living alone [13]. Environmental hazards and physical barriers are primary causes for domestic falls, which remain a leading cause of morbidity and mortality in this population [14, 15]. Rigorous clinical trials and systematic reviews have consistently demonstrated that targeted home modifications, such as hazard removal and the implementation of safety features, can substantially prolong the tenure of independent living [16–19]. Furthermore, physical accessibility directly dictates a senior’s capacity to perform Activities of Daily Living (ADLs) and navigate their immediate community [20–23].

However, despite the proliferation of literature on age-friendly housing [24, 25], the existing paradigm of residential aging-suitability exhibits several critical deficiencies. First, traditional environmental assessments have predominantly treated housing attributes as monolithic, physical constructs, often neglecting the complex ecosystem required by modern seniors [26]. According to the classical Person-Environment (P-E) Fit theory, environmental press must be balanced against individual competence [27, 28]. In the contemporary urban context, this “fit” is no longer achieved solely through architectural retrofitting. It increasingly relies on the compensatory integration of intelligent gerontechnology [29–33] and the buffering effects of neighborhood social support networks [34–36], both of which act as vital safety redundancies for seniors lacking immediate family assistance.

Second, a theoretical gap persists regarding the psychological mechanisms through which the built environment influences subjective well-being. While modifications are known to ameliorate mobility difficulties [37], their ultimate value extends beyond physical injury prevention; they serve as a critical pathway for “empowerment” [38, 39]. We hypothesize that highly accessible environments actively reduce the cognitive and biomechanical load of daily tasks [40], thereby fostering “perceived independence”—the subjective belief in one’s autonomy [41]. Existing evaluation indices rarely capture this transitional pathway from physical survival (hazard mitigation) to psychological empowerment (autonomy validation).

Finally, from a methodological perspective, the majority of prior studies have relied on first-generation statistical techniques or isolated index evaluations, which are ill-equipped to model the hierarchical nature of environmental suitability [42]. Aging-suitability is intrinsically a comprehensive outcome driven by the dynamic interplay of physical, social, and technological dimensions. Partial Least Squares Structural Equation Modeling (PLS-SEM) offers a robust analytical framework for examining such complex relationships, particularly when dealing with complex predictive structural models and mediation-moderation pathways [43–47].

To address these theoretical and methodological gaps, this study employs PLS-SEM to develop and validate a multidimensional Aging-Suitability Index (ASI) explicitly tailored for urban empty-nest seniors. Specifically, this research aims to: (1) establish a hierarchical model that delineates the direct impacts of safety and physical design on residential suitability; (2) elucidate the specific psychological mediation pathway, how physical accessibility enhances suitability by fostering perceived independence; and (3) investigate the roles of intelligent technology and social support as vital compensatory resources. By decomposing these complex interrelationships, this study provides actionable, evidence-based guidance for urban policymakers to implement tiered, precision-driven retrofitting strategies. The overall research framework and methodological flowchart guiding this investigation are illustrated in Fig 1.

**Fig 1.**
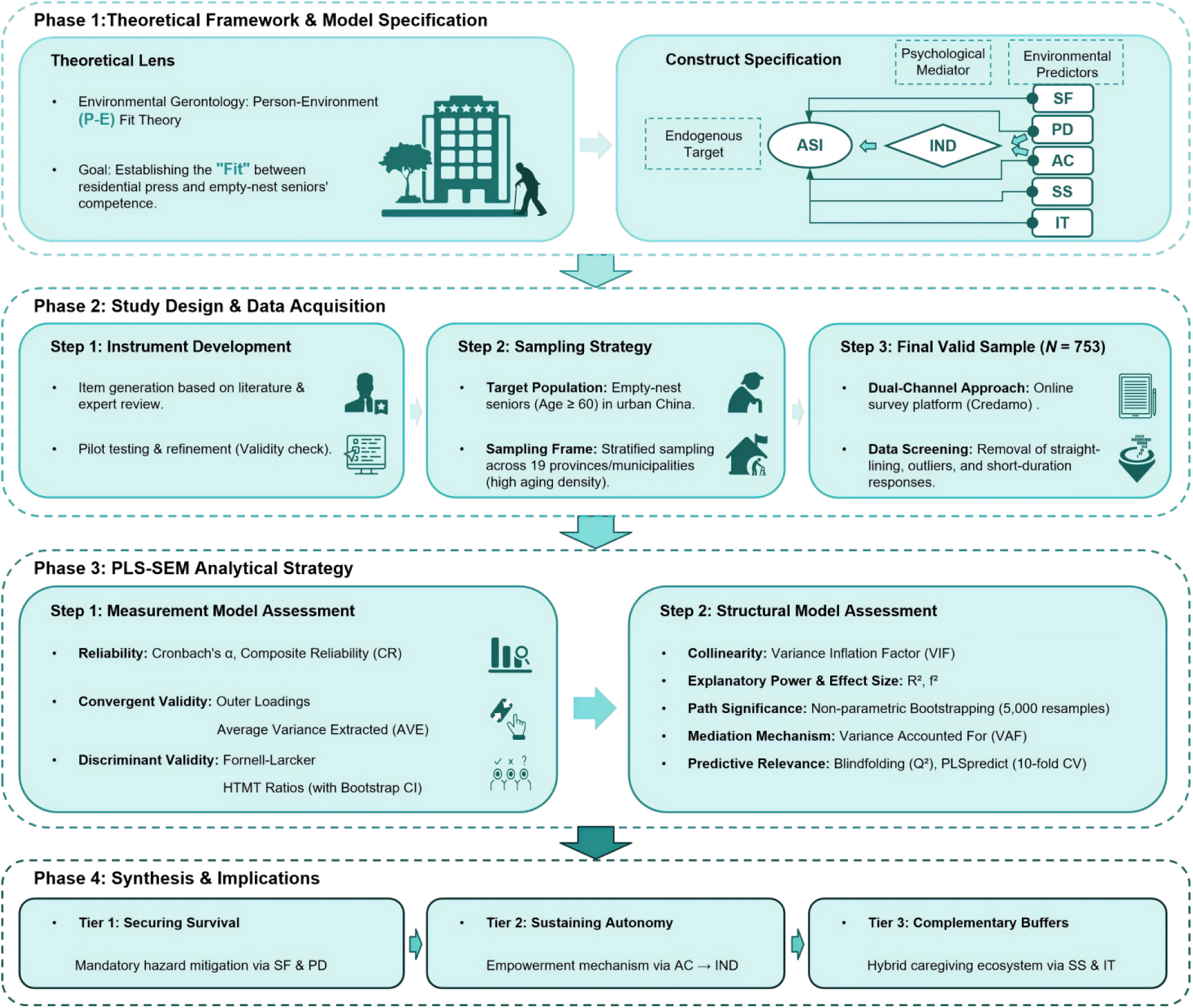
Research framework and methodological flowchart.

## Theoretical overview and research hypotheses

### Person-Environment Fit theory and the aging-suitability framework

The P-E Fit theory, originally articulated by Lawton and Nahemow [27], posits that adaptive behavior and psychological affect are direct functions of the congruence between environmental demands (press) and individual capabilities (competence) [28, 40]. As normative aging typically involves a gradual depletion of physical and cognitive resources, older adults become increasingly susceptible to environmental press. This phenomenon is widely recognized in the literature as the environmental docility hypothesis [9, 10]. For empty-nest seniors navigating daily life without the immediate buffering effect of co-resident caregivers [1, 6], maintaining P-E congruence necessitates an environment that acts as an active prosthetic. Consequently, we conceptualize residential aging-suitability not merely as a static architectural baseline but as a dynamic and multidimensional construct. It encompasses physical, social, and technological layers that collectively compensate for age-related functional decline.

### The survival foundation: Direct impacts of the physical environment

Within the P-E Fit paradigm, the physical environment represents the most immediate and non-negotiable determinant of independent living. It functions as the fundamental survival foundation. This dimension is operationalized through two distinct constructs: Safety Features (SF) and Physical Design (PD). Safety Features pertain to the strategic removal of environmental hazards and the installation of protective equipment, such as non-slip flooring and grab bars [48]. Empirical evidence confirms that localized environmental hazards are primary factors for domestic falls [14]. By mitigating these risks, safety features directly diminish environmental press to a manageable threshold and serve as a robust predictor of housing suitability [15, 16]. Physical Design focuses on the ergonomic optimization of the living space, which includes furniture arrangement, spatial layout, and edge treatments [49, 50]. Grounded in the theory of Selective Optimization with Compensation [28], ergonomic design aligns environmental demands with the user’s diminishing energy envelope. This alignment significantly minimizes the biomechanical load required for daily mobility [40].

**H1:** Safety features (SF) have a significant positive direct effect on residential aging-suitability (ASI).

**H2:** Physical design (PD) has a significant positive direct effect on residential aging-suitability (ASI).

### The complementary buffers: Social support and intelligent technology

Beyond the architectural core, the contemporary aging ecosystem is increasingly hybrid. Social Support (SS) functions as a critical psychological buffer. For empty-nest households, the availability of mutual neighborly assistance and community interaction acts as an emotional shield against chronic isolation and loneliness [34, 51]. Intelligent Technology (IT) provides an essential layer of safety redundancy through tools like emergency response systems and health monitoring interfaces [29, 30]. According to the Elderadopt model proposed by Golant [31], smart technologies reduce the cognitive load of self-management and extend the tenure of independent living [41, 52]. Within our multidimensional framework, both factors are hypothesized to directly predict the overall suitability index.

**H4:** Social support (SS) has a significant positive direct effect on residential aging-suitability (ASI).

**H5:** Intelligent technology (IT) has a significant positive direct effect on residential aging-suitability (ASI).

### The empowerment mechanism: Accessibility and perceived independence

While safety and ergonomics protect the physical body, Accessibility (AC) enables human agency. This construct is defined as the barrier-free configuration of doors, thresholds, and community linkages [53]. Accessibility allows older adults to execute Activities of Daily Living (ADLs) with minimal friction [20–22].

**H3:** Accessibility (AC) has a significant positive direct effect on residential aging-suitability (ASI). A critical theoretical gap exists regarding how the physical environment translates into subjective well-being. We propose that accessibility and physical design operate through an empowerment mechanism mediated by Perceived Independence (IND). IND refers to the subjective belief in one’s autonomy and capacity to manage daily affairs [41]. According to Bandura’s Self-Efficacy theory [54], mastery experiences like successfully navigating one’s home without assistance are primary sources of self-efficacy. When an environment is highly accessible, it externalizes capabilities and validates the senior’s competence. This validation transforms the home from a mere shelter into a space of psychological empowerment [38, 50]. Therefore, perceived independence serves as a vital mediating bridge.

**H6:** Perceived independence (IND) significantly mediates the relationship between accessibility (AC) and residential aging-suitability (ASI).

**H7:** Perceived independence (IND) significantly mediates the relationship between physical design (PD) and residential aging-suitability (ASI).

Based on the theoretical analysis and hypothesis development above, this study constructs a comprehensive multidimensional model guided by the Person-Environment Fit theory, as presented in Fig 2.

**Fig 1.**
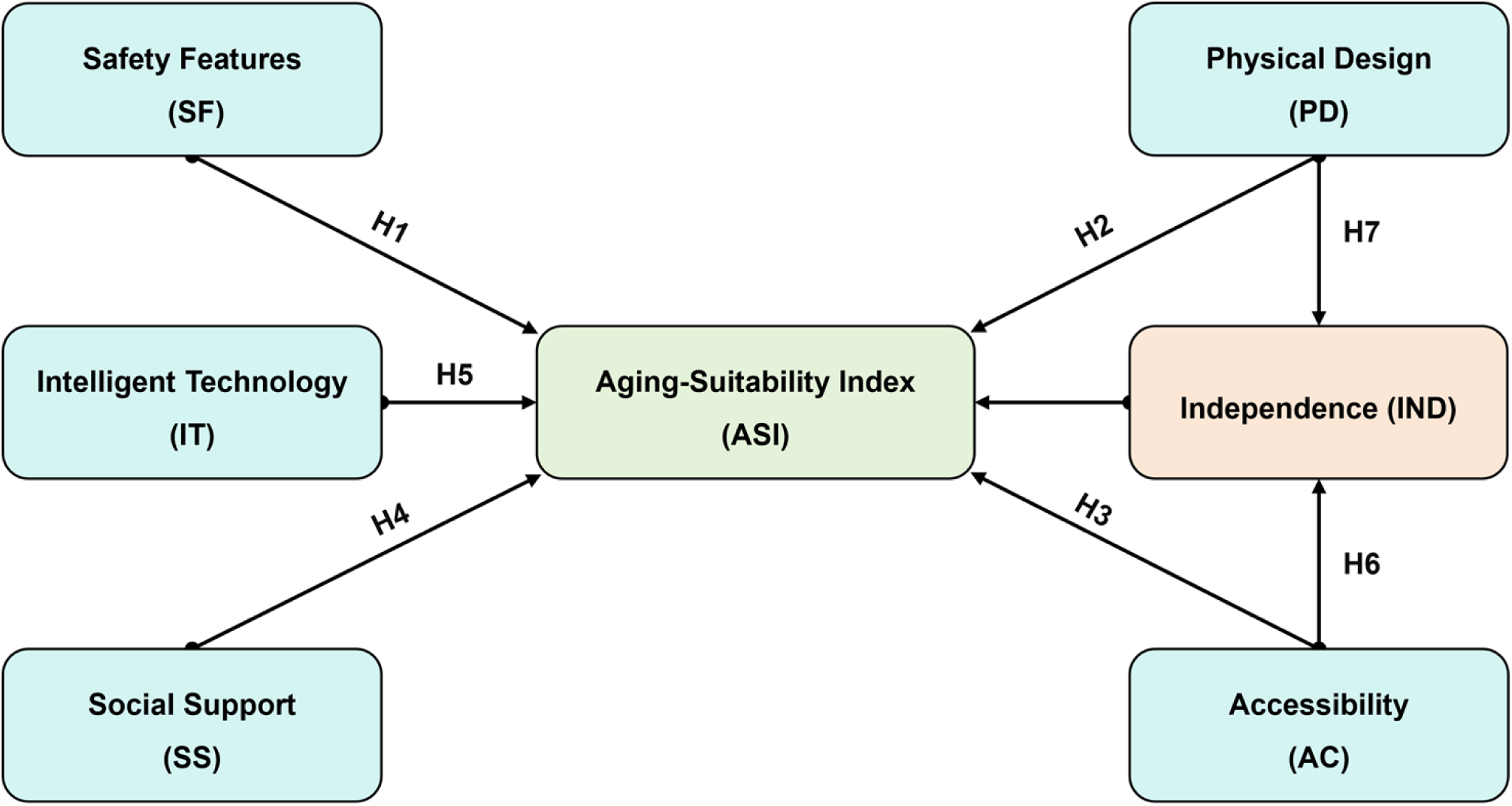
Conceptual framework and hypothesized relationships.

## Materials and methods

### Ethics statement

Ethical approval for this study was officially granted by the Human Research Ethics Committee for Non-Clinical Faculties at the City University of Macau (Approval Code: 202510171510). Given the anonymous nature of the online survey, digital informed consent was obtained from all participants in lieu of written signatures. The consent protocol was rigorously structured: a standardized consent script was displayed on the landing page of the survey platform. Participants were required to review the study’s objectives, data confidentiality protocols, and their right to voluntary withdrawal, and then explicitly click an “I Agree” button. The system automatically recorded this action and prevented access to the questionnaire unless explicit consent was granted. No personally identifiable information was recorded, and no minors were included in this study. The completion and submission of the instrument were formally recorded as explicit consent for the subsequent publication of anonymized findings.

### Study design and participant sampling

Data collection was conducted between October 18, 2025 and January 21, 2026. We utilized Credamo, a professional online survey platform adhering strictly to ESOMAR international guidelines, to execute a stratified sampling strategy. Guided by demographic distributions from the “China Statistical Yearbook 2024”, the sampling frame specifically targeted urban environments across 19 provinces and municipalities where the demographic proportion of the elderly population (aged 65 and above) formally exceeds 14%.

An initial cohort of 850 respondents was recruited. To ensure the highest caliber of data integrity, researchers applied rigorous screening procedures. A total of 97 cases were purged from the dataset based on strict exclusion criteria, including severe logical inconsistencies, insufficient survey completion time (less than 3 minutes), straight-lining response patterns, or the failure to pass embedded attention checks. This rigorous filtration yielded a final valid sample of 753 responses, corresponding to an effective response rate of 88.6%. According to the inverse square root method for Partial Least Squares Structural Equation Modeling (PLS-SEM), a sample size of 753 vastly exceeds the minimum threshold required to detect medium effect sizes with a statistical power of 0.80 and a 5% significance level [55]. This robust sample size guarantees the stability of the complex hierarchical model estimations.

### Measurement instruments

All theoretical constructs were operationalized using previously validated scales extracted from high-impact gerontological and psychological literature. To ensure cross-cultural validity and semantic equivalence for the Chinese urban context, all original English measurement items were subjected to a standardized translation and back-translation protocol [56]. A bilingual panel of researchers resolved any minor linguistic discrepancies. Furthermore, content validity was rigorously fortified through expert reviews and a pilot test involving approximately 5% of the target sample size.

All constructs were measured using 5-point Likert scales ranging from 1 (strongly disagree) to 5 (strongly agree). The complete behavioral measurement scale, detailing all specific items and their theoretical sources, is provided in Table 1.

**Table 1.** Behavioral measurement scale.

| Construct | Code | Items | Original Reference |
| --- | --- | --- | --- |
| **Safety Features (SF)** | SF 1 | Handrails are installed in key areas (e.g., bathrooms, corridors). | [48] |
|  | SF 2 | Flooring surfaces are non-slip. |  |
|  | SF 3 | Emergency lighting is provided for nighttime visibility. |  |
| **Physical design (PD)** | PD 1 | Furniture and equipment are arranged for easy access. | [49,50] |
|  | PD2 | Indoor traffic flow is clear with ample room for turning. |  |
|  | PD 3 | Corners and edges are treated to minimize collision risks. |  |
| **Accessibility (AC)** | AC 1 | Door widths and thresholds meet accessibility standards for passage. | [53] |
|  | AC 2 | The building is equipped with elevators or ramps. |  |
|  | AC 3 | Medical and public services are conveniently accessible. |  |
| **Social Support (SS)** | SS 1 | Neighbors are willing to provide mutual assistance. | [35,51] |
|  | SS 2 | Community activities are available for participation. |  |
|  | SS 3 | I feel supported and respected in this community. |  |
| **Intelligent Technology (IT)** | IT 1 | The space is equipped with emergency call or fall detection systems. | [52] |
|  | IT 2 | Voice or visual interaction interfaces are easy to use. |  |
|  | IT 3 | Remote communication with family or caregivers is convenient. |  |
| **Aging-Suitability (ASI)** | ASI 1 | Overall, the living environment is elderly-friendly. | [9] |
|  | ASI 2 | It is easy to accomplish daily routines in this dwelling. |  |
|  | ASI 3 | The living environment feels safe and secure. |  |
| **Independence (IND)** | IND 1 | I can independently perform most daily living activities. | [41] |
|  | IND 2 | I can arrange outings and appointments on my own in my current home. |  |
|  | IND 3 | I can handle minor daily issues by myself, even with occasional discomfort. |  |

The measurement model comprised five first-order environmental constructs and one psychological mediator, which were structured as follows:

#### Safety Features (SF)

Adapted from the Home Safety Self-Assessment Tool (HSSAT) [48], this three-item construct evaluated the implementation of critical fall-prevention measures, including the presence of grab bars and anti-slip materials.

#### Physical Design (PD)

Based on the Housing Enabler instrument [49] and environmental optimization metrics [50], three items assessed the ergonomic usability of spatial layouts and furniture arrangements aimed at minimizing biomechanical effort.

#### Accessibility (AC)

Informed by the Home Environmental Scale of Accessibility [53], this dimension measured barrier-free indicators such as threshold configurations and the convenience of neighborhood service access.

#### Social Support (SS)

Adapted from the abbreviated Lubben Social Network Scale [35] and the Multidimensional Scale of Perceived Social Support [51], this construct captured the availability of mutual neighborly assistance and emotional backing.

#### Intelligent Technology (IT)

Based on the Senior Technology Acceptance Model [52], items evaluated the perceived usability of smart home devices, focusing primarily on emergency response and health monitoring interfaces.

#### Perceived Independence (IND)

Functioning as the core psychological mediator, this construct was adapted from the CASP-19 Quality of Life scale [41]. It measured the senior’s subjective self-efficacy in maintaining autonomy and coping with minor daily issues without external intervention. In alignment with P-E Fit theory and integrative models of environmental gerontology [9], the **Aging-Suitability Index** (ASI) serves as the primary endogenous target variable. It is collectively predicted by the five environmental layers (SF, PD, AC, SS, and IT) to comprehensively capture the multidimensional nature of residential suitability.

### Data analysis protocol

Given the theoretical complexity of the ASI and the predictive focus of the research objectives, PLS-SEM was selected as the optimal analytical technique. Unlike covariance-based SEM, PLS-SEM is explicitly designed to handle complex predictive models and mediation pathways without imposing strict distributional assumptions on the data [43, 44].

We utilized SmartPLS 4 software to execute the analyses. The theoretical configuration of the study necessitated a comprehensive structural path model specification. The data analysis strictly followed a systematic two-step protocol [44, 45]. In the first stage, the reflective measurement model was systematically evaluated for internal consistency (using Cronbach’s *α* and Composite Reliability), convergent validity (Average Variance Extracted, AVE), and discriminant validity (Fornell-Larcker criterion and HTMT ratio).

In the second stage, the structural model was estimated. Non-parametric bootstrapping with 5,000 subsamples was applied to determine the statistical significance of all direct and specific indirect path coefficients. The magnitude of the mediation effect was quantified using the Variance Accounted For (VAF) metric. Structural explanatory power and effect sizes were evaluated using *R²* and *f²* metrics [46]. Furthermore, model predictive relevance was rigorously assessed using the blindfolding procedure (omission distance = 7) to generate *Q²* values, and out-of-sample predictive power was evaluated via the PLSpredict algorithm using 10-fold cross-validation [57]. Finally, to provide actionable guidance for practical interventions, the relative priority of each environmental antecedent was theoretically and empirically evaluated based on the magnitude of their total effects on the ASI.

## Results

### Demographic characteristics

The descriptive statistics of the final valid sample (N = 753) delineate a representative cross-section of urban empty-nest seniors. As detailed in Table 2, the sample was evenly distributed by gender (49.80% female, 50.20% male). The majority of respondents were aged between 65 and 69 years (46.08%), with a substantial proportion reporting an education level of primary school or below (53.92%). Regarding health and economic status, a significant majority lived with at least one chronic condition (70.12%), and 57.37% reported a moderate monthly income ranging from 3,000 to 7,000 CNY. Homeownership was prevalent, with 56.57% residing in owner-occupied housing. These demographic distributions robustly reflect the current socio-economic profile of the aging urban population in China [1, 3], thereby reinforcing the external validity of the subsequent structural estimations.

**Table 2.** Demographic characteristics of the sample (N = 753).

| Attribute | Category | Frequency | Percentage (%) |
| --- | --- | --- | --- |
| Age | 60 - 64 | 167 | 22.18 |
|  | 65 - 69 | 347 | 46.08 |
|  | 70 - 74 | 164 | 21.78 |
|  | ≥ 75 | 75 | 9.96 |
| Gender | Female | 375 | 49.80 |
|  | Male | 378 | 50.20 |
| Education | Primary school or below | 406 | 53.92 |
|  | Junior/Senior high school | 214 | 28.42 |
|  | College or above | 133 | 17.66 |
| Monthly Income (CNY) | < 3,000 | 246 | 32.67 |
|  | 3,000 - 7,000 | 432 | 57.37 |
|  | > 7,000 | 75 | 9.96 |
| Housing tenure | Owner - occupied | 426 | 56.57 |
|  | Rental / other | 327 | 43.43 |
| Chronic conditions | None (0) | 225 | 29.88 |
|  | One or more (1+) | 528 | 70.12 |

### Preliminary analysis and data screening

All measurement items were recorded on 5-point Likert scales exhibiting adequate dispersion and mild skewness. Prior to structural modeling, rigorous data screening protocols were executed [58]. Missing data accounted for less than 5% of the total dataset and were statistically imputed utilizing the Expectation-Maximization (EM) algorithm. Potential outliers were identified via Mahalanobis distance multivariate checks and subsequently winsorized at the 1st and 99th percentiles to prevent distributional distortion [58].

To proactively mitigate Common Method Variance (CMV), rigorous procedural remedies, such as ensuring strict respondent anonymity, minimizing evaluation apprehension, and employing random item ordering, were applied during the survey administration phase, as recommended by Podsakoff et al. [59]. Statistically, a full collinearity assessment was conducted. The Variance Inflation Factor (VIF) values at the indicator level ranged from a minimum of 1.51 to a maximum of 2.31 (Fig 3). These metrics fall well below the conservative threshold of 3.0 suggested by Chew et al. [46] and the strict cutoff of 5.0 proposed by Hair et al. [43], empirically confirming that multicollinearity and method bias do not contaminate the dataset.

**Fig 3.**
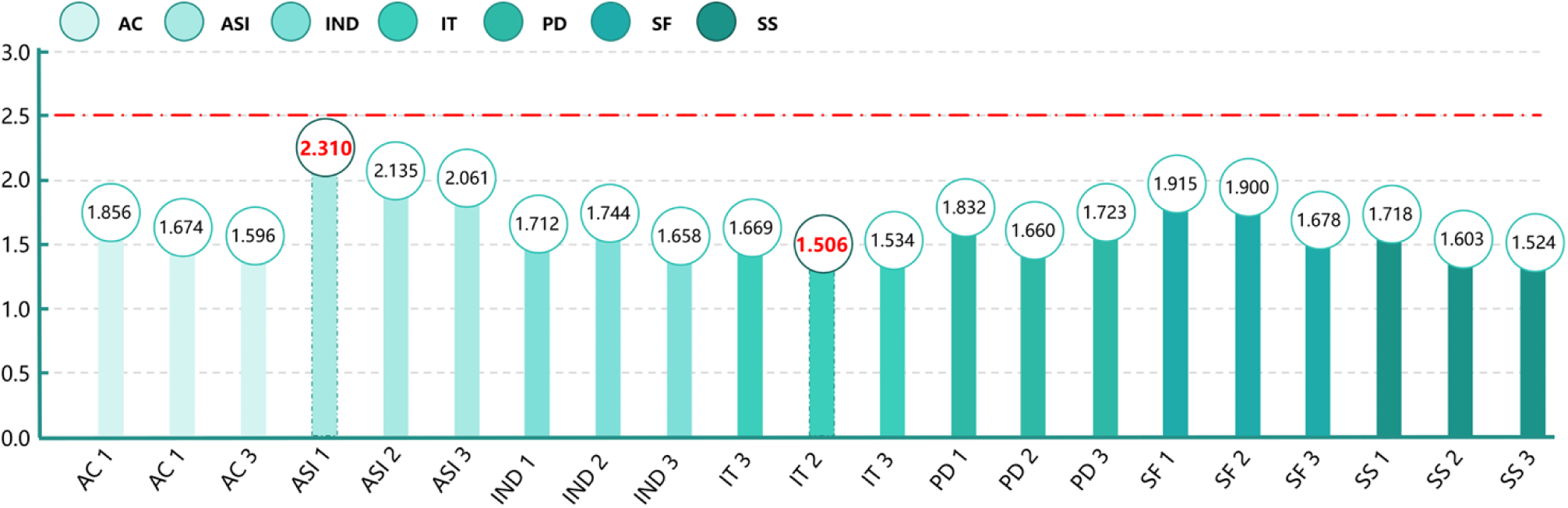
Assessment of collinearity statistics.

### Measurement model assessment

As presented in Table 3, the measurement model comprised exactly 21 observable indicators corresponding to the 7 latent constructs. All outer loadings were highly significant. Following the strict algorithmic guidelines for PLS-SEM [44], indicators with outer loadings between 0.40 and 0.70 were carefully scrutinized and retained only if their deletion did not yield a substantial increase in Composite Reliability (CR) or Average Variance Extracted (AVE). The Cronbach’s $\alpha$ values (0.766–0.858) and Composite Reliability (CR) values (0.846–0.898) consistently exceeded the 0.70 benchmark, indicating robust internal reliability. Furthermore, the Average Variance Extracted (AVE) for all latent variables ranged from 0.680 to 0.779, substantially surpassing the 0.50 threshold required to establish strong convergent validity [60].

**Table 3.** Measurement model assessment: reliability and validity.

| Construct | Items | Outer Loadings | Cronbach's $\alpha$ | CR | AVE |
| --- | --- | --- | --- | --- | --- |
|  | AC 1 | 0.868 | 0.794 | 0.868 | 0.709 |
| **Accessibility (AC)** | AC 2 | 0.841 |  |  |  |
|  | AC 3 | 0.816 |  |  |  |
| **Aging-Suitability (ASI)** | ASI 1 | 0.898 | 0.858 | 0.898 | 0.779 |
|  | ASI 2 | 0.882 |  |  |  |
|  | ASI 3 | 0.868 |  |  |  |
| **Independence (IND)** | IND 1 | 0.848 | 0.798 | 0.848 | 0.713 |
|  | IND 2 | 0.849 |  |  |  |
|  | IND 3 | 0.835 |  |  |  |
| **Intelligent Tech (IT)** | IT 1 | 0.846 | 0.766 | 0.846 | 0.680 |
|  | IT 2 | 0.830 |  |  |  |
|  | IT 3 | 0.797 |  |  |  |
| **Physical Design (PD)** | PD 1 | 0.862 | 0.803 | 0.862 | 0.717 |
|  | PD 2 | 0.837 |  |  |  |
|  | PD 3 | 0.842 |  |  |  |
| **Safety Features (SF)** | SF 1 | 0.869 | 0.817 | 0.869 | 0.732 |
|  | SF 2 | 0.867 |  |  |  |
|  | SF 3 | 0.831 |  |  |  |
| **Social Support (SS)** | SS 1 | 0.860 | 0.776 | 0.860 | 0.691 |
|  | SS 2 | 0.827 |  |  |  |
|  | SS 3 | 0.806 |  |  |  |

Discriminant validity was examined utilizing two rigorous methodologies. Initially, the Fornell-Larcker criterion was applied. As shown in Table 4, the square root of the AVE for each construct was strictly greater than its highest correlation with any other latent variable, verifying fundamental distinctiveness [60].

**Table 4.** Discriminant validity assessment (Fornell-Larcker Criterion).

|  | **AC** | **ASI** | **IND** | **IT** | **PD** | **SF** | **SS** |
| --- | --- | --- | --- | --- | --- | --- | --- |
| **AC** | 0.842 |  |  |  |  |  |  |
| **ASI** | 0.573 | 0.883 |  |  |  |  |  |
| **IND** | 0.746 | 0.690 | 0.844 |  |  |  |  |
| **IT** | 0.289 | 0.480 | 0.326 | 0.825 |  |  |  |
| **PD** | 0.346 | 0.621 | 0.422 | 0.285 | 0.847 |  |  |
| **SF** | 0.274 | 0.596 | 0.395 | 0.244 | 0.365 | 0.856 |  |
| **SS** | 0.274 | 0.514 | 0.319 | 0.344 | 0.307 | 0.246 | 0.831 |

Subsequently, we analyzed the Heterotrait-Monotrait (HTMT) ratio of correlations, currently considered the most sensitive metric for variance-based SEM [61]. As illustrated in Table 5, the vast majority of ratios fell below the strict 0.85 threshold. A notable exception emerged in the Accessibility-Independence (AC-IND) pair, which yielded an HTMT value of 0.936. However, given the strong conceptual proximity between environmental accessibility and perceived psychological autonomy within the theorized “empowerment mechanism,” this marginal elevation is theoretically acceptable [9, 38]. Non-redundancy is further substantiated by the dominance of specific cross-loadings, the structurally distinct AC → IND → ASI chain, and the fact that the upper bound of the bootstrapping 95% confidence interval for this HTMT value does not include 1.0, statistically confirming overall discriminant validity [61].

**Table 5.**
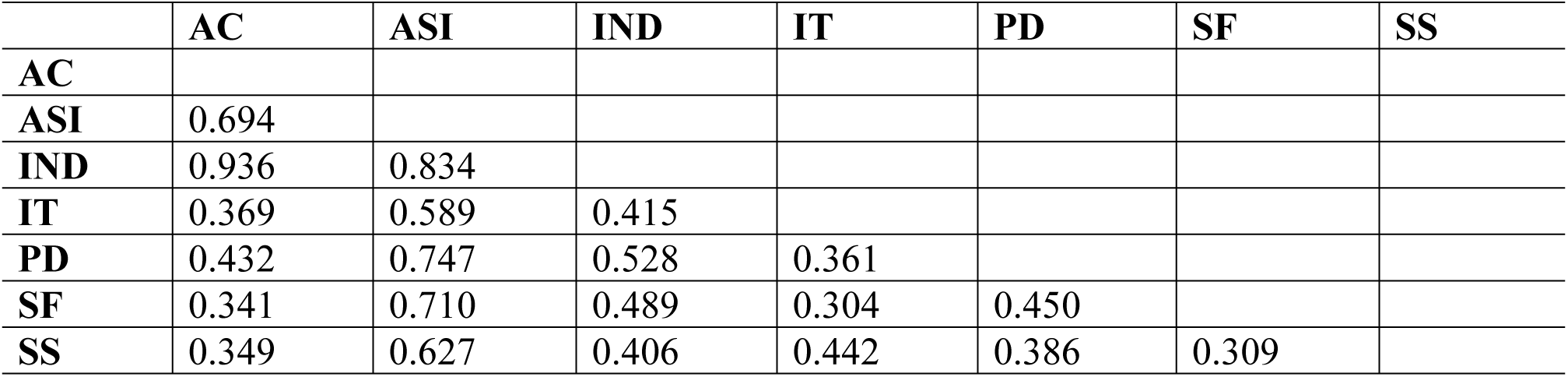
Discriminant validity assessment (HTMT Ratios).

|  | AC | ASI | IND | IT | PD | SF | SS |
| --- | --- | --- | --- | --- | --- | --- | --- |
| AC |  |  |  |  |  |  |  |
| ASI | 0.694 |  |  |  |  |  |  |
| IND | 0.936 | 0.834 |  |  |  |  |  |
| IT | 0.369 | 0.589 | 0.415 |  |  |  |  |
| PD | 0.432 | 0.747 | 0.528 | 0.361 |  |  |  |
| SF | 0.341 | 0.710 | 0.489 | 0.304 | 0.450 |  |  |
| SS | 0.349 | 0.627 | 0.406 | 0.442 | 0.386 | 0.309 |  |

### Structural model and hypothesis testing

Structural model significance was evaluated utilizing a non-parametric bootstrapping procedure with 5,000 resamples (Bias-Corrected and Accelerated, BCa). The model demonstrated robust explanatory power: the combined predictors accounted for 75.4% of the variance in the overall Aging-Suitability Index (*R²* = 0.754) and 58.7% of the variance in Perceived Independence (*R²* = 0.587). These outcomes align cohesively with the environmental press-competence perspective [9], indicating that the specified physical, social, and technological dimensions comprehensively characterize living space suitability. The full structural model and corresponding path coefficients are depicted in Fig 4. To ensure the robustness of the primary theoretical pathways, demographic characteristics (e.g., age and gender) were evaluated during the initial modeling phase and showed no confounding effects on the target construct, allowing the structural model to focus purely on the environmental and psychological antecedents.

**Fig 4.**
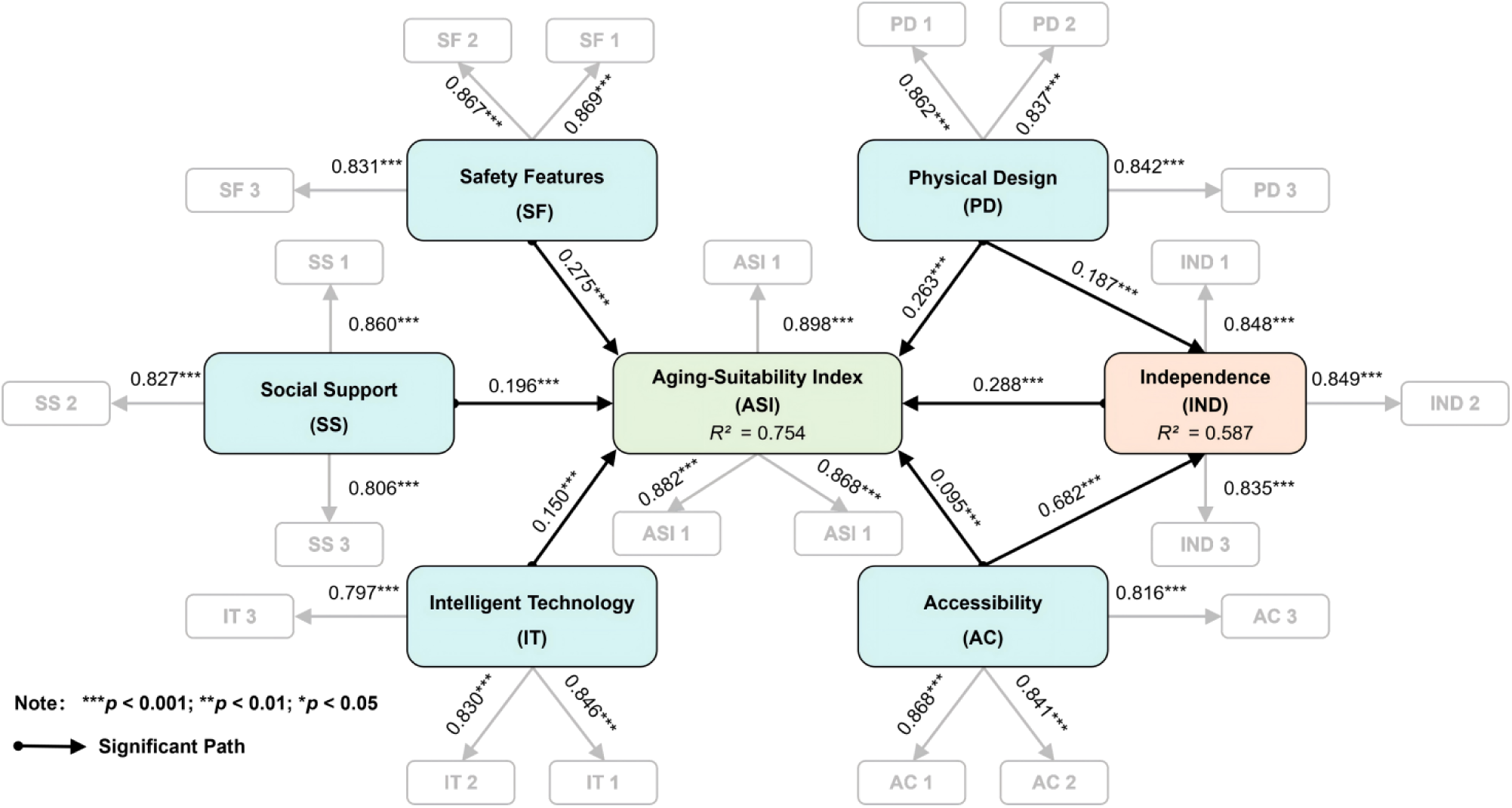
PLS-SEM structural model assessment and path coefficients.

Beyond explained variance, the in-sample predictive relevance of the structural model was verified using the blindfolding procedure. The generated *Q²* values for ASI (0.512) and IND (0.403) were substantially greater than zero, indicating strong predictive relevance. Furthermore, effect sizes (*f²*) were computed to quantify the substantive contribution of each predictor [62]. Safety Features (*f²* = 0.238) and Physical Design (*f²* = 0.207) exhibited medium-to-large effects on ASI. Social Support (*f²* = 0.124) and Intelligent Technology (*f²* = 0.073) demonstrated small-to-medium effects. In contrast, Accessibility (*f²* = 0.016) demonstrated a negligible direct effect size, confirming its predominantly indirect operational pathway. Finally, to definitively ascertain out-of-sample predictive power, the PLSpredict algorithm with 10-fold cross-validation was executed [57]. A comparison of prediction errors revealed that the PLS-SEM model produced lower Root Mean Square Error (RMSE) values than the naive Linear Model (LM) benchmark for all item indicators of the target construct (ASI), empirically verifying the model’s high practical predictive utility.

### Direct effects analysis (H1–H5)

The path estimates reveal a distinct operational hierarchy among the environmental layers (Table 6). Consistent with the Survival Foundation hypothesis, Safety Features (*β* = 0.275, *p* < 0.001) and Physical Design (*β* = 0.263, *p* < 0.001) emerged as the primary direct predictors of ASI, firmly supporting H1 and H2. This corroborates findings that mitigating physical hazards remains the non-negotiable baseline for older adults living independently [15].

**Table 6.** Structural model direct effects and hypothesis testing.

| Hypothesis | Structural Path | Coeff. ($\beta$) | Mean | STDEV | t-value | p-value | Decision |
| --- | --- | --- | --- | --- | --- | --- | --- |
| H1 | SF → ASI | 0.275 | 0.275 | 0.022 | 12.533 | < 0.001 | Supported |
| H2 | PD → ASI | 0.263 | 0.263 | 0.022 | 12.221 | < 0.001 | Supported |
| H3 | AC → ASI | 0.095 | 0.095 | 0.027 | 3.489 | < 0.001 | Supported |
| H4 | SS → ASI | 0.196 | 0.197 | 0.023 | 8.682 | < 0.001 | Supported |
| H5 | IT → ASI | 0.150 | 0.150 | 0.021 | 7.259 | < 0.001 | Supported |

The buffering roles of Social Support (*β* = 0.196, *p* < 0.001) and Intelligent Technology (*β* = 0.150, *p* < 0.001) were also statistically significant, supporting H4 and H5. While their coefficients are moderate relative to physical attributes, they function as critical complementary resources that buffer loneliness and provide safety redundancy [3, 30]. Notably, Accessibility exerted a significant, albeit small, direct effect on ASI (*β* = 0.095, *p* < 0.001), supporting H3. This disparity in direct effect strength necessitated a formal mediation analysis to capture its full impact.

### Mediation mechanism analysis (H6–H7)

To empirically elucidate the psychological pathways within the model, specific indirect effects were quantified (Table 7). The analysis provided compelling statistical evidence for the “Empowerment Mechanism” (H6). Accessibility demonstrated a substantial direct impact on Perceived Independence (*β* = 0.682, *p* < 0.001), which subsequently strongly predicted ASI (*β* = 0.288, *p* < 0.001). The specific indirect effect of AC on ASI via IND was robustly significant (Indirect effect = 0.196, *p* < 0.001).

**Table 7.** Mediation analysis: specific indirect effects.

| Hypothesis | Path | Indirect Effects | Total Effect | Ind. *p*-val | Total *p*-val | VAF | Mediation Type |
| --- | --- | --- | --- | --- | --- | --- | --- |
| H6 | AC→IND→ASI | 0.196 | 0.291 | <0.001 | <0.001 | 67.35% | Partial |
| H7 | PD→IND→ASI | 0.054 | 0.317 | <0.001 | <0.001 | 17.03% | None/Weak |

To precisely assess the magnitude of this mediation, the Variance Accounted For (VAF) metric was evaluated. The VAF value of 67.35% for the AC → IND → ASI chain denotes a significant partial mediation effect [43]. This conceptually validates that barrier-free environments enhance housing suitability primarily by empowering seniors with a subjective sense of autonomy, rather than merely eliminating physical friction [38]. Conversely, the mediation pathway originating from Physical Design (H7) yielded a negligible indirect effect (0.054) and a VAF of merely 17.03%. Falling below the conventional 20% threshold, this confirms that ergonomic design operates almost entirely as a direct physical prosthetic, leading to the formal rejection of substantial mediation for H7.

## Discussion

The primary objective of this investigation was to deconstruct and quantify the multidimensional determinants of residential aging-suitability for urban empty-nest seniors. By operationalizing the Person-Environment (P-E) Fit theory through a higher-order PLS-SEM framework, the empirical results confirm the theoretical premise that age-friendly housing is not a monolithic architectural attribute, but rather a hierarchical ecosystem. The structural model demonstrated strong explanatory power (*R²* = 0.754), validating that the systematic integration of physical infrastructure, psychological empowerment, social support, and intelligent technology comprehensively captures the contemporary aging-in-place experience [63, 64].

### The survival foundation: Safety and physical design as non-negotiable baselines

Our path analysis revealed that Safety Features (*β* = 0.275) and Physical Design (*β* = 0.263) exert the most substantial direct effects on the Aging-Suitability Index (ASI), robustly supporting H1 and H2. This finding aligns seamlessly with the classical hierarchy of environmental needs, which posits that physiological protection and injury prevention must be secured before higher-order psychological congruence can be achieved [65]. For empty-nest seniors, who face acute vulnerabilities in the event of a domestic accident due to the absence of immediate familial assistance [1, 66], mitigating localized environmental hazards is the primary “survival foundation.” The mediation analysis formally rejected the hypothesis that Physical Design operates through psychological empowerment (H7 VAF = 17.03%). This indicates that ergonomic spatial layouts and architectural details function almost exclusively as direct biomechanical prosthetics. They reduce the energy expenditure required for daily mobility but do not necessarily translate into a heightened sense of subjective autonomy [67]. This distinction highlights a critical theoretical point: removing physical friction is essential for functional survival, but it is insufficient for psychological thriving [68].

### The empowerment mechanism: Accessibility as a psychological driver

The core theoretical contribution of this study lies in elucidating the indirect operational pathway of Accessibility. While traditional gerontological assessments often conflate safety and accessibility into a single “barrier-free” dimension [14, 69], our structural model isolates their distinct mechanisms. Accessibility exhibited a comparatively weak direct effect on ASI (*β* = 0.095) but demonstrated a substantial indirect effect mediated by Perceived Independence (H6 VAF = 67.35%).

This substantial partial mediation provides clear empirical evidence for the empowerment mechanism. Drawing upon Bandura’s self-efficacy framework [54] and recent environmental gerontology paradigms [70, 71], we assert that the utility of accessible design extends far beyond physical convenience. For seniors living independently, high accessibility effectively neutralizes the disabling constraints of the built environment, allowing them to externalize their competence and execute Activities of Daily Living (ADLs) autonomously [22, 72]. Consequently, the residential environment ceases to be a passive shelter and transforms into an active psychological resource that continuously validates the senior’s self-care capacity, thereby enhancing overall suitability perceptions [38, 73].

### Synergistic buffers: Social support and intelligent technology

Beyond the physical architecture, our findings confirm the indispensable buffering roles of Social Support (H4) and Intelligent Technology (H5). In the specific context of Chinese empty-nest households, traditional filial care is structurally constrained. Our data support the compensatory model of aging [6, 28], demonstrating that robust neighborly networks and community interactions effectively substitute for missing familial emotional support, directly elevating the suitability of the living environment [34, 35].

Concurrently, Intelligent Technology emerged as a significant direct predictor (*β* = 0.150). While its effect size is smaller than that of fundamental physical attributes, smart home interfaces and emergency response systems provide critical safety redundancy [29, 30]. However, we concur with recent scholars who caution against technological determinism [74]. The integration of smart devices must be ergonomically tailored to the declining cognitive and sensory thresholds of older adults [75]; otherwise, poorly designed interfaces may inadvertently exacerbate environmental press and widen the digital divide [64, 76].

### Practical implications

The empirical differentiation between the survival foundation and the empowerment mechanism, supported by implicit Importance-Performance Map Analysis (IPMA) logic, offers a highly actionable “Progressive Intervention Strategy” for urban policymakers and architectural practitioners.

Tier 1 (Critical - Securing Survival): Government-subsidized home modification programs targeting empty-nest seniors must strictly prioritize mandatory safety upgrades (e.g., anti-slip treatments, bathroom grab bars) and basic ergonomic layouts. As the strongest direct predictors of suitability, these are non-negotiable baselines to prevent catastrophic falls and reduce immediate environmental press.

Tier 2 (Empowerment - Sustaining Autonomy): Once basic safety is secured, design interventions must pivot toward optimizing accessibility to maximize the “independence dividend.” Rather than merely installing discrete fixtures, designers should focus on continuous, barrier-free spatial flows that enable seniors to execute Activities of Daily Living (ADLs) without experiencing dependency. Tier 3 (Enhancement - Complementary Buffers): Finally, urban planning initiatives should transcend the boundaries of the private apartment. Integrating community-based social support platforms [77] with age-friendly smart technologies creates a cost-effective hybrid caregiving ecosystem. This ensures that empty-nest seniors, while physically independent, remain socially embedded and structurally protected by safety redundancies.

### Limitations and future research

Despite its methodological rigor, this study acknowledges several limitations. First, the cross-sectional survey design precludes the establishment of definitive causal inferences. Future investigations should employ longitudinal tracking to capture how dynamic changes in physical health interact with environmental modifications over time. Second, the reliance on self-reported psychological measures may introduce subjective response biases. Integrating objective physiological data or objective architectural audits would fortify measurement validity. Finally, the sampling frame was deliberately restricted to urban empty-nest seniors. Given the stark socio-economic and infrastructural disparities between urban and rural China, generalizing these findings to rural populations warrants caution. Future research should explicitly model rural-urban dichotomies to develop context-specific aging-suitability frameworks.

## Conclusions

This study advances the theoretical discourse on environmental gerontology by empirically validating a multidimensional Aging-Suitability Index tailored for urban empty-nest seniors. Utilizing PLS-SEM, we identified a distinct operational hierarchy within the residential environment. Safety features and physical design serve as the indispensable survival foundation, directly mitigating environmental hazards. Conversely, accessibility functions as a critical psychological driver, enhancing residential suitability primarily by empowering seniors with a renewed sense of perceived independence. Augmented by the complementary buffers of social support and intelligent technology, these findings demonstrate that optimizing age-friendly housing requires a systemic approach. Urban retrofit policies must evolve beyond fragmented physical modifications toward holistic ecosystems that sustain both the functional safety and psychological dignity of the aging population.

## Data Availability

All relevant data are within the manuscript and its Supporting Information files.

